# Understanding and comparing the globe-trotting cancer patient with the locally managed patient: A case control study

**DOI:** 10.1101/2022.04.13.22273435

**Authors:** Mary W Wangai, Frederick K Wangai, Francis Njiiri, John Kinuthia, Enan N. Wangai, Catherine Nyongesa, Paul Wangai

## Abstract

Medical tourism is characterized by people seeking treatment abroad for various medical conditions due to varied reasons, many of whom benefit from specialized care for non-communicable diseases. Conversely, there are associated negative effects such as medical complications and weakened health systems. Currently, there is paucity of scientific evidence on factors influencing seeking treatment benefits abroad. This study sought to compare patient-related factors associated with choice of cancer treatment center locally or abroad, to understand reasons for seeking treatment outside Kenya.

**Materials and Methods:** As a case-control study, 254 cancer patients were randomly sampled to compare responses from those who chose to receive treatment abroad or in Kenya. The cases were recruited from Ministry of Health while the controls from Kenyatta National Hospital and Texas Cancer Center. Data was analyzed using SPSS Software Version 21. Descriptive statistics, bivariate and multiple logistic regression analysis was carried out. Level of significance was set at 5%.

**Results:** Out of 254 respondents, 174 (69.5%) were treated for cancer in Kenya and 80 (31.5%) in India. We found that cost effectiveness was a significant factor for over 73% of all respondents. The study revealed independent predictors for seeking treatment in India were: monthly income higher than US$ 250; every additional month from diagnosis increased likelihood by 1.16 times; physician advice (Odds Ratio(OR) 66; 95% Confidence Interval(CI) 7.9 −552.9); friends and family (OR 42; 95% CI 7.07-248.6); and perception of better quality of care (OR 22.5; 95% CI 2.2-230.6).

**Conclusion:** Reasons patients with cancer sought treatment in India are multifactorial. Several of these can be addressed to reverse out-ward bound medical tourism and position Kenya to be a regional hub as per the country’s development blueprint. It will require strengthening the health system accordingly and sensitizing the medical fraternity and general public on the same.

## Introduction

Medical tourism is an emerging and rapidly growing industry globally, in both developing and developed countries. The term refers to clients or patients who specifically traveling out of their country of origin (outward bound) or into a destination country to receive health care (inward bound) [1–3]. Most people seek treatment abroad for management of non-communicable diseases, like cancer. The Kenyan Ministry of Health reports also show that cancer is the 3^rd^ highest cause of mortality [4] and the commonest health condition prevalent among medical tourists. Despite the benefits of medical tourism, the industry can negatively impact health systems in both source and destination countries to the disadvantage of vulnerable populations [2,5–7]. Additionally, some patients acquire medical complications and multidrug resistant microorganisms [1,8–17].

The reasons for the rapid growth of the industry are complex, multifactorial and augmented by globalization and reduced cost of travel. Often times the reasons are due to the fact that home healthcare systems may be inadequate, unavailable, unaffordable or proscribed [18]. Crooks et al categorize the motivation for travel into 3, namely: procedure based; cost-based and travel-based [19]. Medical tourism is also considered an attractive health commodity of high economic value by governments, middle men, facilitating entities and health facilities. Globally, its current market turnover is 24-40 billion US dollars (US$), with potential to generate US$ 28.0 billion by the end of 2024 [20]. The potentially high economic value attached to medical tourism has caused several developing and developed countries to focus on developing this industry [21], Kenya included [22,23]. These countries need to have concerted efforts to increase funding to build and strengthen their healthcare systems to manage the potential influx of patients seeking specialized healthcare as well as for their citizenry [24]. For Kenya, this implies that even with a percent growth domestic product of 5% in 2019 [25], the government expenditure on health will have to increase from its stagnated position of between 6 and 9.2% [24,26] to the intended 12% and above [24,27].

Outward bound medical tourism is growing rapidly in Kenya, like in other developing countries, although it has not been well documented or scientifically researched. This study sought to identify patient-related factors that influence outward-bound travel for cancer treatment, compared with those who seek treatment in country, and thus contribute evidence-based recommendations for reversing the ‘tide’.

## Materials and Methods

### Research Objective

To compare factors that influence patients’ choice of cancer treatment centers, located outside Kenya or those in the country, specifically at Kenyatta National Hospital’s Cancer Treatment Center (KNH) and Texas Cancer Center (TCC) in Nairobi, Kenya.

### Study Design

This is a case-control study that explored the patient related factors that influence choice of Cancer Treatment Centre, abroad or in-country

### Target and Study Population

The case group comprised patients who obtained travel approval for cancer treatment abroad from Ministry of Health (MOH) and returned back into the country after the first cycle/round of therapy. The control group comprised patients who underwent the first cycle/round of treatment within the country at either KNH or TCC in Nairobi. Parents of minors provided the required data concerning their minors since they are the primary decision makers in relation to selecting the country in which to obtain treatment.

### Study Sites

The sites were selected because they formed central points of congregation of target populations in large enough to allow for random selection of study respondents, and included:

a. **The Ministry of Health (MOH)** mandated to deal with health policy and regulation provides approvals for requests for treatment abroad which are needed to facilitate financial support from the National Health Insurance Fund. In the devolved system of Government, the Ministry oversees the national referral health facilities in line with the Constitution 2010 [28];
b. **The Kenyatta National Hospital (KNH)** which is the largest public referral hospital in Kenya, offers subsidized comprehensive treatment for cancer. It receives patients from all the health facilities in the 47 Counties. In 2019, the Centre had a high patient workload with 23,985 visits.
c. **The Texas Cancer Centre** is a medium cost private comprehensive cancer treatment hospital that had a total workload of 15,214 patient visits in 2019. It was selected to be part of the study because of its high workload and the centrality of the site for management of cancer patients.

### Sample size

A minimal sample size of 216 respondents (72 study subjects per site) was determined using the formula described by Fleiss et al [29], taking into account equal ratio of cases to controls, desired power of 80%, 20% non-response rate and 5% level of significance,.

### Data Collection tools

The structured questionnaire was administered to eligible respondents who met the study objectives by research assistants. Although most of the questions were not open ended, the tool included 11 instances where respondents were given opportunity to specify any other alternative answer.

The data collection tool was based on literature from key peer-reviewed journals[7,19,30–33]. The data elements included in the tool included socio-demographic characteristics, disease profile and medical treatment, treatment financing, factors influencing choice of health facility and country, perception of quality of care at chosen facility, and chosen treatment center and destination country.

We also noted there is lack of consensus on a standard definition of who a medical tourist is and a therefore a lack of globally agreed-upon methods of data collection [7].

### Data Collection Procedures

At the MOH, a research assistant perused records of cancer patients who received approval for treatment abroad, while at KNH and TCC patient files were used to select those who fulfilled the inclusion criteria. At all sites, these records were serialized and subjected to random selection using a computer based program. The selected eligible study subjects were called, informed about the study and requested to participate. Thereafter appointments for consenting and researcher-assisted data collection were made with those that agreed to participate. Face-to-face or telephone interviews were conducted to collect the required data after consent were provided. The parents or guardians of minors participated on behalf of their children as they were the primary decision makers, and requested to provide written consent. Upon completion of the questionnaire, the forms were scanned for completeness and accuracy before storing them safely in a secure room.

### Data Management

Data from the structured questionnaires was cleaned manually and then electronically using Statistical Package for the Social Sciences (SPSS) software version 21. Data analysis was carried out using the same software. Descriptive analysis was done using frequencies and cross tabulation to determine level of significance on the all variables. Measures of central tendency and dispersion were determined for continuous variables. Logistic regression models were used to identify independent predictors of country choice for cancer treatment we conducted bivariate analysis for each possible influencing factor on both cases and controls, followed by a multivariate regression for all factors that showed significant association with choice of country (primary outcome). Odds ratios and corresponding 95% confidence intervals(CI) were documented for the influencing factors in choice of country to show strength of association. Statistical tests were performed at 5% (P< 0.05) level of significance.

### Ethical Approval

The study was reviewed and approved by the Kenyatta National Hospital/University of Nairobi (KNH/UON) Ethics and Research Committee (ERC) in August 2018.

## Results

### Sociodemographic characteristics

A total of 254 patients were enrolled into the study with 174 (68.5%) seeking cancer treatment in Kenya at the Kenyatta National Hospital(KNH) and Texas Cancer Center(TCC) and 80 (31.5%) abroad from MOH records. All the study respondents who sought treatment abroad selected India as their country of choice. The study showed the mean age of respondents was 50 years (standard deviation(SD) 15.84), with 159(63.4%) being over 45 years of age (see Table 1). Nearly two-thirds, 167 (65.7%) of respondents were female, and at least 205 (80%) had primary school education (Table 1).

**Table 1:** Socio-demographic characteristics of the sampled study subjects.

| Variables | Categories | Number | Percent (%) |
| --- | --- | --- | --- |
| Gender | Male | 87 | 34.25 |
|  | Female | 167 | 65.7 |
| Age groups (years) | <14 | 5 | 2.0 |
|  | 15-29 | 24 | 9.6 |
|  | 30-44 | 63 | 25.1 |
|  | 45-59 | 88 | 35.1 |
|  | 60-74 | 57 | 22.7 |
|  | >75 | 14 | 5.6 |
| **Marital Status** | Never married | 26 | 10.3 |
|  | Currently married | 197 | 77.9 |
|  | Separated or Divorced | 8 | 3.2 |
|  | Widowed | 15 | 5.9 |
|  | Declined to answer | 7 | 2.8 |
| **Location** | Urban area | 97 | 38.3 |
|  | Rural area | 156 | 61.7 |
| **Education background** | No formal schooling | 15 | 5.9 |
|  | Primary school | 80 | 31.5 |
|  | Secondary/ High School | 83 | 32.7 |
|  | College/ University | 42 | 16.5 |
| **Occupation background** | Government employee | 28 | 11.0 |
|  | Non-governmental employee | 29 | 11.4 |
|  | Unemployed | 82 | 32.3 |
|  | Self-employed | 77 | 30.3 |
|  | Farmer | 31 | 12.2 |

Nearly two-thirds of the respondents, 165 (65%), were employed or had income generating activities. Out of these, 80 of them earned a median monthly salary of about US$ 390 (minimum US$ 7 to maximum US$ 4,000) (Table 4). The length of time (duration) the study subjects knew they suffered from cancer ranged from 1 to 188 months, with a median time of 13 months (IQR 4-17 months).

The study respondents suffered various forms of cancers; reproductive tract cancers (126 cases, 49.6%), gastrointestinal tract cancers 46 (18.1%), blood related malignancies 30 (11.8%), and Ear, Nose and Throat cancers 24 (9.4%) (Figure 1).

**Figure 1:**
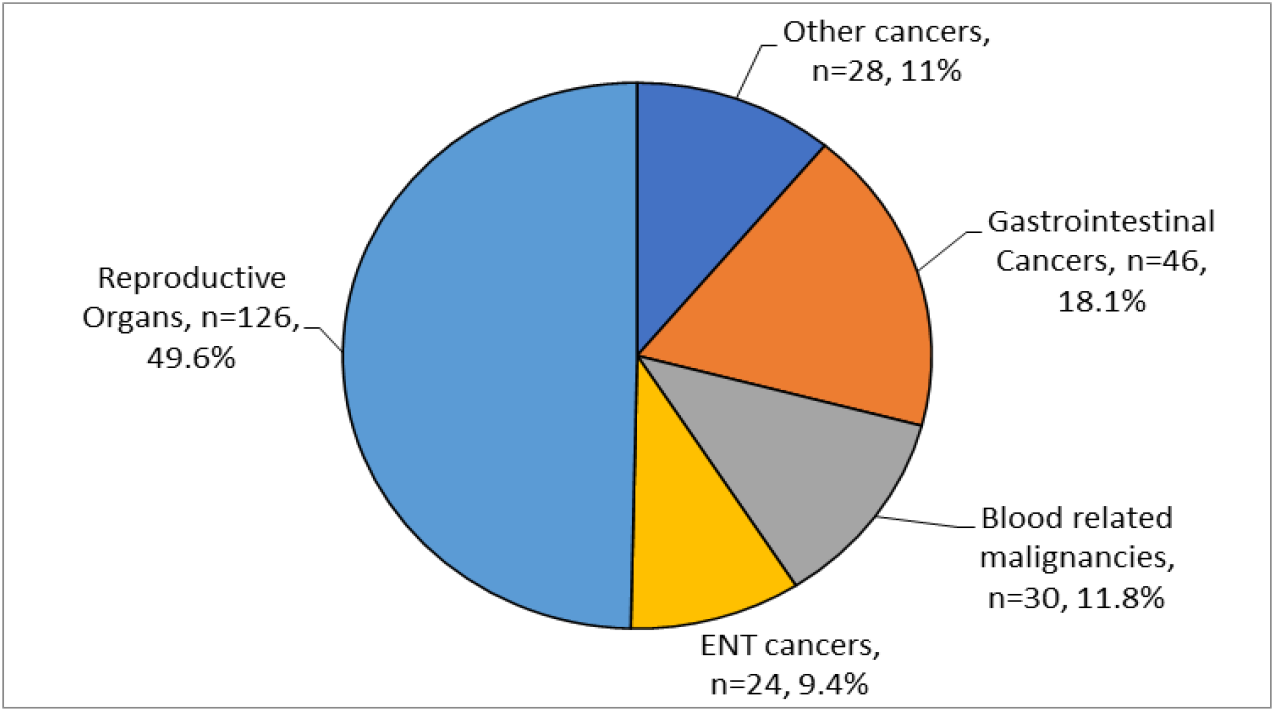
Types of Cancers in Categories.

The specific cancers respondents had were breast 59 (23.2%), cervix 50 (19.7%), oesophagus 16 (6.3%), prostate 10 (3.9%) and other types of cancers 119 (46.9%). One hundred and two (60%) participants had various laboratory tests, while 70 (28%) underwent a positron emission tomography (PET) scanning, and 40 (16%) respondents had other radiological tests performed on them. Study respondents were treated using chemotherapy, 221 (87.0%), radiotherapy, 72 (28%), surgery 53 (21.0%), 10 (4%) bone marrow transplantation and 2 respondents received brachytherapy.

### Reasons Given for Choice of Country to Obtain Cancer Treatment

The majority, 210 (83%), of respondents indicated that advice from their physicians influenced their choice of country in which to seek treatment. Other key influencing factors included; cost-effectiveness (n=185, 73%), advice from friends and family (n=77, 70%), perceived access to highly specialized health care services (n=164 64.8%), perceived access to highly skilled and experienced health workers (n=131 52%) and quality of care (n=177, 70%) (Table 2 below).

**Table 2:** Reasons Given for Choice of Country for Cancer Treatment.

| Factor | Number | Percent |
| --- | --- | --- |
| Physicians advice | 210 | 83.0 |
| Cost effectiveness | 185 | 73.1 |
| Friends and family | 177 | 70.0 |
| Quality of care | 177 | 70.0 |
| Perceived access to specialized health care services | 164 | 64.8 |
| *Reputation of the Country | 156 | 61.9 |
| Perceived access to highly skilled and experienced health workers | 131 | 52.0 |
| Other reasons | 2 | 50.0 |
| Combine treatment with sight-seeing | 60 | 23.7 |
| Combine treatment with business | 45 | 17.8 |

*\*Perception that the Country has the reputation to offer the required specialized cancer treatment health services*

### Sociodemographic Characteristics and Their influence on Choice of Country

Bivariate analysis, using chi-square statistics, revealed significant differences between the cases and controls. With regard to the sociodemographic characteristics, we found that traveling to India was associated with the male gender (X^2^=5.4, p=0.021), urban dwelling (X^2^=83.3, p<0.0001), higher education background (X^2^=105.9, p<0.0001) and government or NGO employment (X^2^=72.3, p<0.0001) (Table 3). We found that there was a significant association between occupation and choice of country. Of those that were treated in India, 22 (27.5%) were government employees, 18 (22%) were self-employed, 20 (9.7%) were retirees or unemployed (p<0.0001) (Table 3).

**Table 3:**
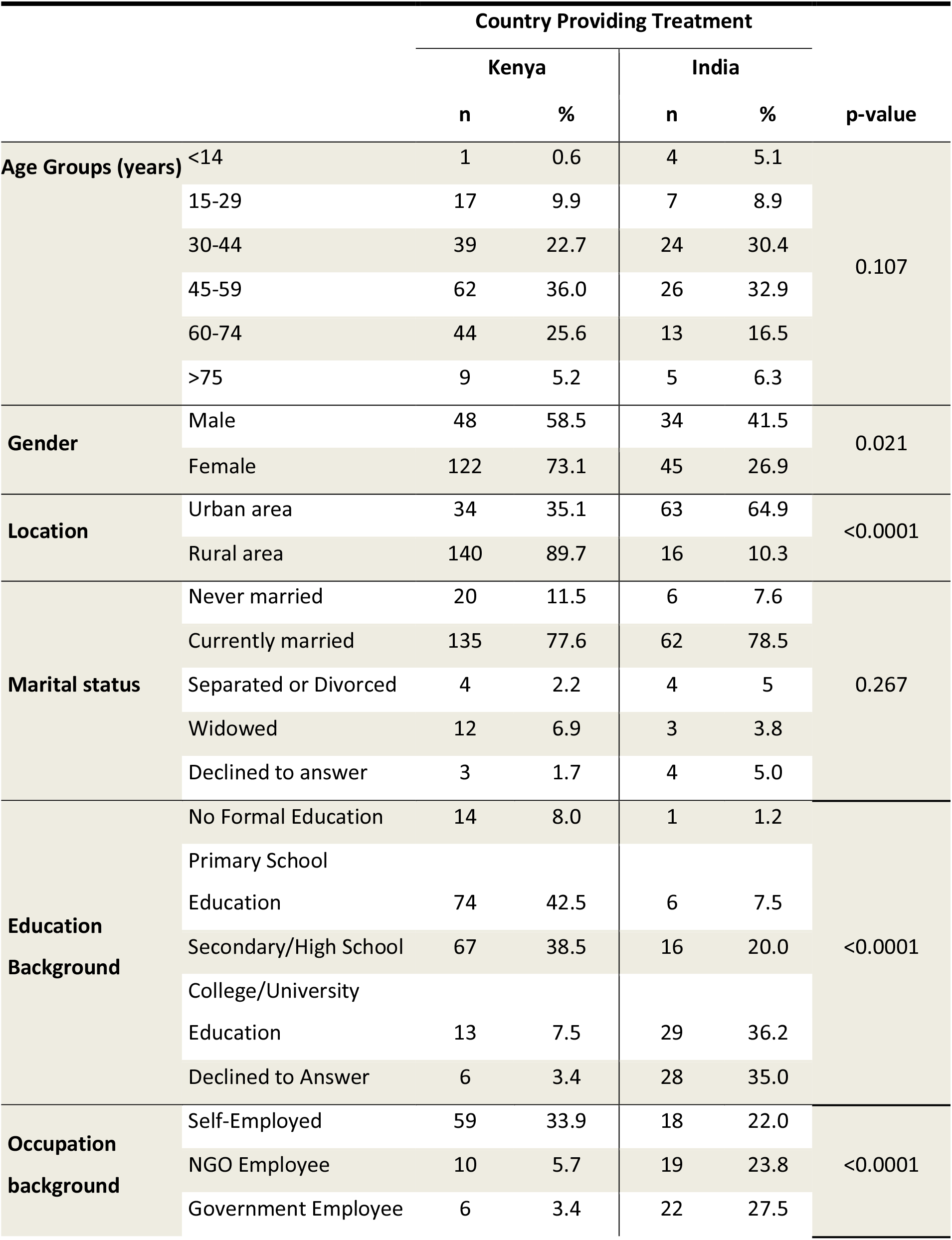
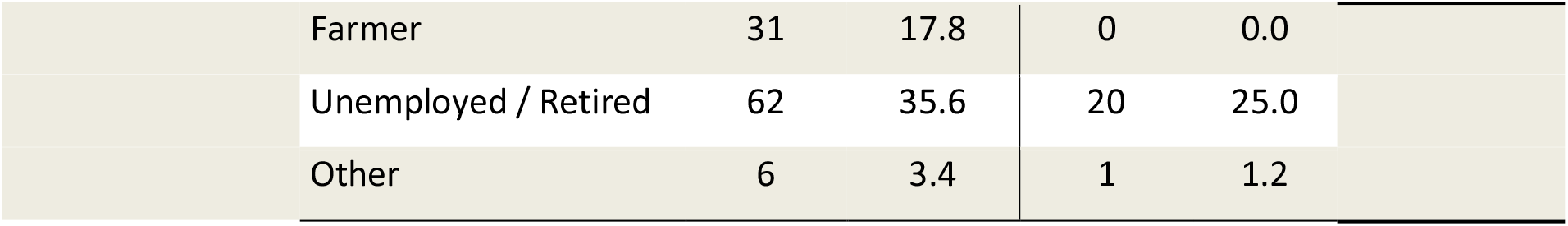
Associations between Sociodemographic characteristics and Choice of Country.

|  |  | Country Providing Treatment |  |  |  | p-value |
| --- | --- | --- | --- | --- | --- | --- |
|  |  | Kenya |  | India |  |  |
|  |  | n | % | n | % |  |
| Age Groups (years) | <14 | 1 | 0.6 | 4 | 5.1 | 0.107 |
|  | 15-29 | 17 | 9.9 | 7 | 8.9 |  |
|  | 30-44 | 39 | 22.7 | 24 | 30.4 |  |
|  | 45-59 | 62 | 36.0 | 26 | 32.9 |  |
|  | 60-74 | 44 | 25.6 | 13 | 16.5 |  |
|  | >75 | 9 | 5.2 | 5 | 6.3 |  |
| Gender | Male | 48 | 58.5 | 34 | 41.5 | 0.021 |
|  | Female | 122 | 73.1 | 45 | 26.9 |  |
| Location | Urban area | 34 | 35.1 | 63 | 64.9 | <0.0001 |
|  | Rural area | 140 | 89.7 | 16 | 10.3 |  |
| Marital status | Never married | 20 | 11.5 | 6 | 7.6 | 0.267 |
|  | Currently married | 135 | 77.6 | 62 | 78.5 |  |
|  | Separated or Divorced | 4 | 2.2 | 4 | 5 |  |
|  | Widowed | 12 | 6.9 | 3 | 3.8 |  |
|  | Declined to answer | 3 | 1.7 | 4 | 5.0 |  |
| Education Background | No Formal Education | 14 | 8.0 | 1 | 1.2 | <0.0001 |
|  | Primary School Education | 74 | 42.5 | 6 | 7.5 |  |
|  | Secondary/High School | 67 | 38.5 | 16 | 20.0 |  |
|  | College/University Education | 13 | 7.5 | 29 | 36.2 |  |
|  | Declined to Answer | 6 | 3.4 | 28 | 35.0 |  |
| Occupation background | Self-Employed | 59 | 33.9 | 18 | 22.0 | <0.0001 |
|  | NGO Employee | 10 | 5.7 | 19 | 23.8 |  |
|  | Government Employee | 6 | 3.4 | 22 | 27.5 |  |

|  |  |  |  |  |  |
| --- | --- | --- | --- | --- | --- |
|  | Farmer | 31 | 17.8 | 0 | 0.0 |
|  | Unemployed / Retired | 62 | 35.6 | 20 | 25.0 |
|  | Other | 6 | 3.4 | 1 | 1.2 |

The study showed that choice to travel to India was significantly associated with higher monthly income (p <0.0001) and longer duration from the time the respondents were diagnosed with cancer (p<0.0001). Study respondents who chose treatment in India had known of their diagnosis for an average period of 26.2 months (SD= 25.9, range 1-188 months), while those treated in Kenya had an average duration of 8 months (SD=10.3, range 1-90 months) (Table 4).

**Table 4:** Factors associated with Choice of Treatment Centre: Income and Knowledge Diagnosis.

|  |  | N | Mean | Std. Deviation | Minimum | Maximum | p-value |
| --- | --- | --- | --- | --- | --- | --- | --- |
| **Monthly income (US\$)** | Kenya | 28 | 136.79 | 123.59 | 7.00 | 500.00 | |
|  | India | 52 | 701.54 | 689.99 | 100.00 | 4000.00 | <0.0001 |
|  | Total | 80 | 503.88 | 621.33 | 7.00 | 4000.00 |  |
| **Duration since diagnosis** | Kenya | 173 | 8.01 | 10.315 | 1 | 90 |  |
|  | India | 78 | 26.23 | 25.911 | 1 | 188 | <0.0001 |
|  | Total | 251 | 13.67 | 18.746 | 1 | 188 |  |

### Types of Cancers, their Management and their Effect on Country of Choice

With regard to cancer categories, there was a statistically significant association between diagnosis of blood related malignancies and provision of cancer treatment in India (X2=74.68; p<0.0001). The majority of respondents, 107 (84.9%) diagnosed with reproductive organ cancers were managed in Kenya (Figure 2).

**Figure 2:**
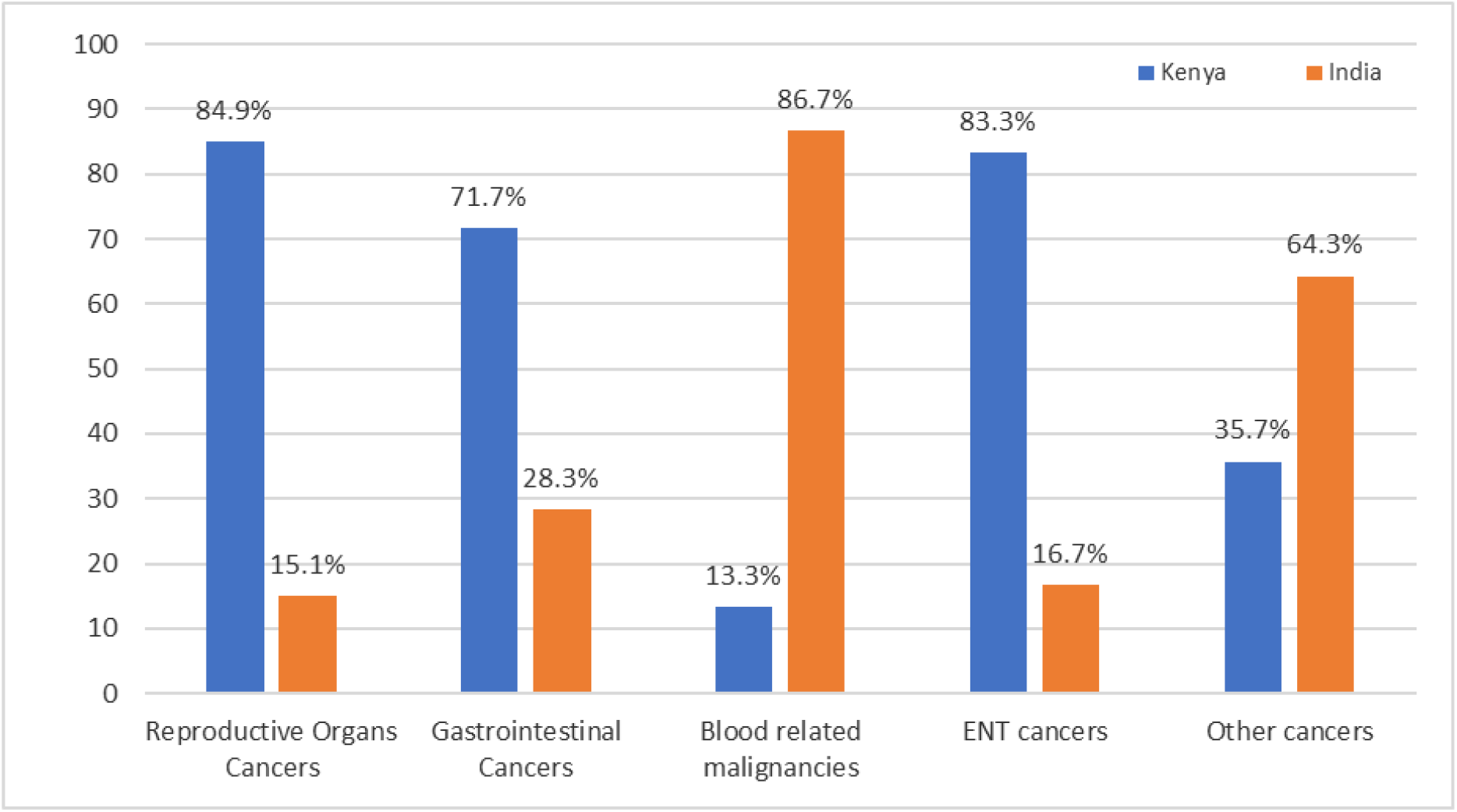
Proportion of Study Participants disaggregated by the Type of Cancers and the Country in which they received treatment.

There was a statistically significant association between the referring health facility and the choice of treatment center (X^2^= 105.6, p=<0.0001). We found that private hospitals referred a higher proportion of respondents to India, 59 (73.8 %), while government hospitals referred only 8 (5.6%) patients.

The study revealed the two main cancer management financers were the NHIF and out-of-pocket house-hold funds; 146 (57.3%) and 140 (55%) respondents, respectively. Private insurance companies funded 7 (3%) of respondents and employers funded 2 (1%) of responders. There was no association between the source of funding for treatment and the country chosen in which to receive treatment (X2= 0.1, p=0.79). The NHIF funded 47(58.8%) of respondents that sought treatment in India while 98 (56.6%) respondents were funded receive treatment in Kenya.

### Factors that influenced Choice of Country for Cancer Treatment

We found a statistically significant association between the country of choice and perception of the country’s capacity to offer required cancer treatment (X^2^= 25.6, p<0.0001), advice from respondent’s physician (X^2^= 5.64, p=0.018), perceived QOC (X^2^= 19.0, p<0.0001), perceived access to specialized health care services (X^2^= 28.5, p<0.0001), opportunity to combine sightseeing (X^2^= 64.9, p<0.0001) amongst others (Figure 3). Sixty-seven (84.8%) of the respondents treated in India were influenced by their perception of the country’s facilities’ capacity to provide cancer treatment. Similarly, 70 (88.6%) were influenced by perceived QOC to be provided, 59 (74.7%) by respondent’s physician and 64 (82.1%) by availability of skilled and experienced health workers (Figure 3).

**Figure 3:**
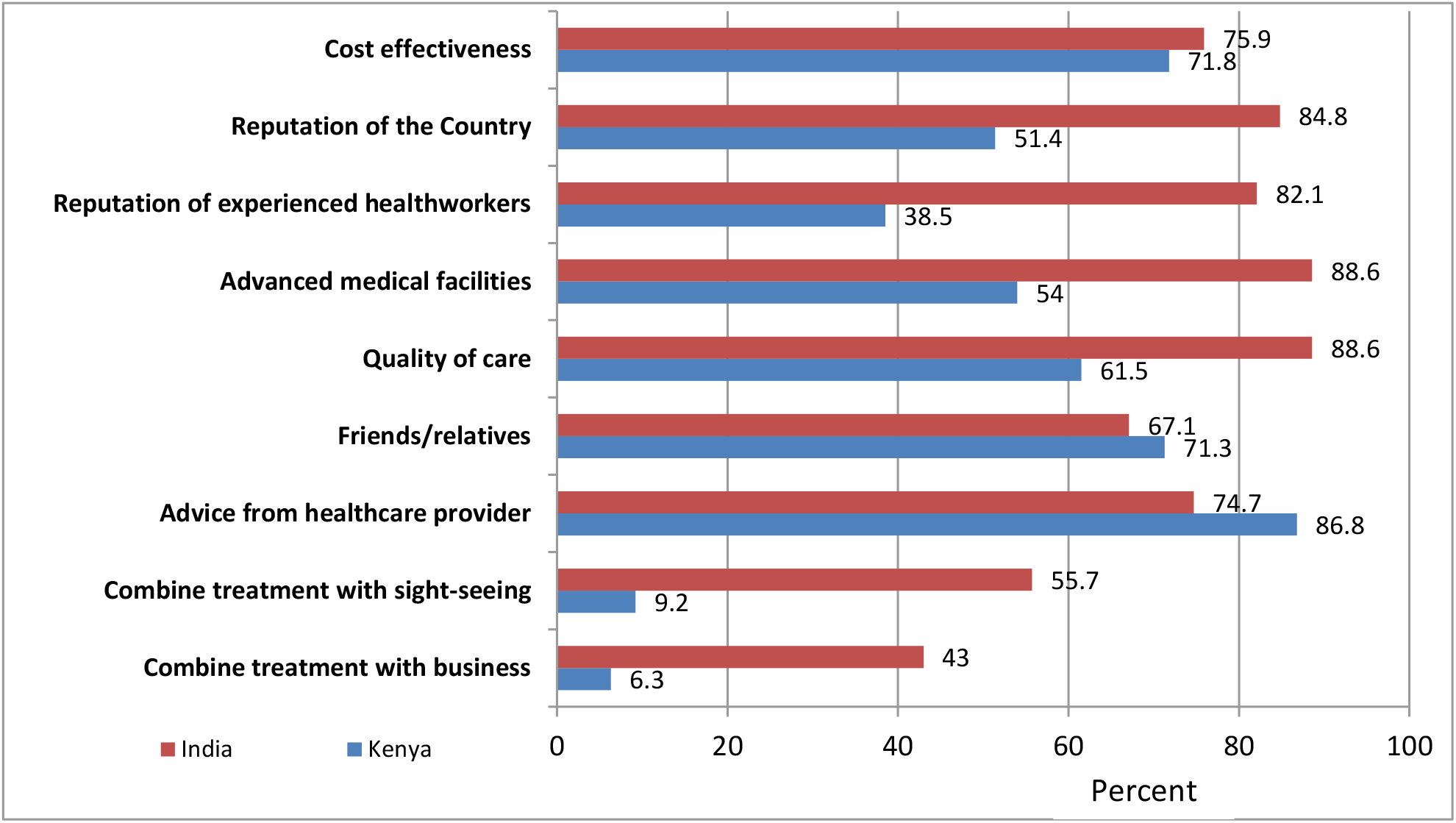
Proportion of Study Respondents and the influencing factors for Choice of Treatment Center. NB: Reputation of the country refers to the respondent’s perception that the Country has the reputation to offer the required specialized cancer treatment health services

### Independent Predicators for Choice Country

Using logistic regression models we identified independent predictors for choice of country. When controlling for gender and other sociodemographic factors, monthly income and duration from diagnosis (in months) were identified as independent predicators. The likelihood for choosing treatment in India was found to be 38.9 times higher for cancer patients who earned US$ 250 monthly and above (p<0.0001, 95%CI 7.5-201.3). Every additional month from diagnosis was associated with increased likelihood of treatment in India by 1.16 times (p= 0.005, 95% CI 1.046-1.28).

When controlling for other factors, we found other significant independent predictors for choice of country. The adjusted odds for seeking treatment in India were 66.2 (95% CI 7.9 −552.9) and 42 (95% CI 7.07-248.6) times higher upon advice from physicians and from friends and family, respectively. Additionally, anticipation to receive better quality of care was another independent predictor, (OR=22.5, 95% CI 2.2-230.6) (Table 5).

**Table 5:**
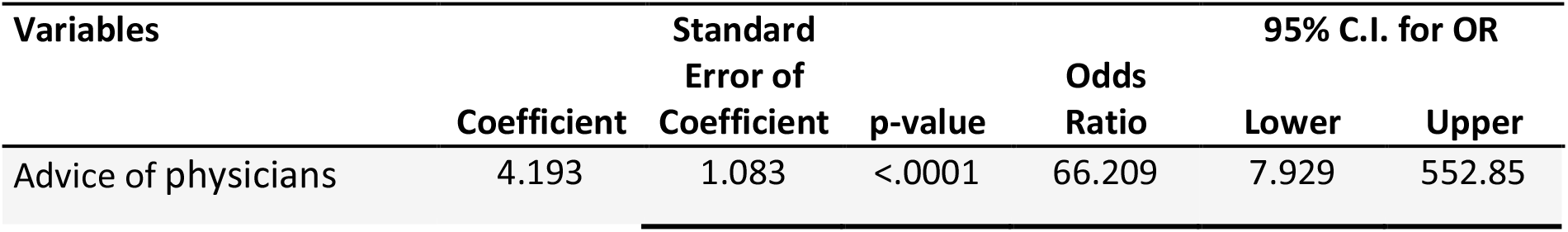
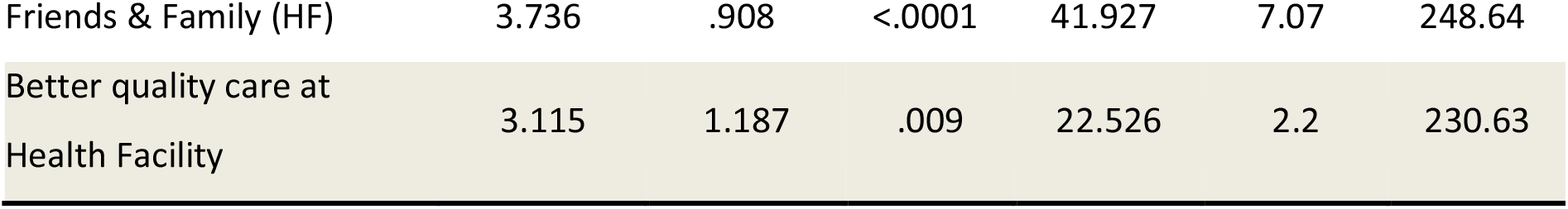
Independent Factors Influencing of selecting Country to Receive Treatment.

## Discussion

There is paucity of systematically generated information about Kenya’s outward-bound medical tourism, yet it is a growing industry locally and globally. This study sought to characterize selected aspects of medical tourism in Kenya by understanding the key patient-related factors that influence choice of treatment out of the country with specific reference to management of cancer.

### Characteristics of Study Subjects and Disease profile

Our study compared 80 study participants who received cancer treatment in abroad, with 174 other participants who chose treatment in Kenya. In this case-control study, cancer patients treated in abroad formed the case group, while those treated in Kenya at KNH and Texas Cancer Center formed our control group. All 80 study respondents treated abroad chose to go to India as their treatment destination of choice. This is was similar to other African patients who travel to Asia, the majority go to India in search of specialized quality, cost-effective health care and customer care that may not be available in their home countries [34].

Much like the Global Cancer Statistics 2020, cancer is a leading cause of mortality in Kenya, and an important barrier to increasing life expectancy [4,35]. Hence cancer deprives the country of its most important productive segment of the population and development resource, the human workforce. The median age of the respondents was 50 years with nearly two-thirds being 45 years and older. Globally a larger proportion of cancer patients (70%) are 50 years and older [35], depicting slightly earlier onset of cancer in the Kenyan population. Most of the commonest cancers in the Globocan Kenya statistics report are similar with those of our respondents, namely; cervix, breast, esophagus, gastrointestinal and prostate cancer. Additionally like Kenya, cancer is the commonest disease for which patients are referred from other southern Sub-Sahara countries into South Africa [37].

Irrespective of the country the respondents chose to receive treatment in, the majority received chemotherapy as the initial mode of therapy. Management of cancer was financed by the National Hospital Insurance Fund (NHIF) or using out-of-pocket household finances, without demonstrable statistically significant difference between those who sought treatment in Kenya or India. This implies that NHIF equally finances treatment in and out of the country. Interestingly, the biggest group that were supported to receive treatment in India by NHIF were government employees (41.9%) and the self-employed (30.4%) respondents. These respondents are among the 19% of the Kenyan population that have health insurance. Unfortunately, the insured are not equitably located across the country. As expected, the wealthy are more likely to have health insurance than the poorest quintile; 42% and 3% respectively [24,27). Thus financing for cancer care in Kenya may come from various sources. Further, anecdotal information reveals that fund raising for health care is often done among friends and relatives as a cultural norm in the country. Other probable sources include selling of assets such as land and livestock.

### Patient Related Factors Associated with Medical Tourism

Bivariant analysis of the socio-demographic factors demonstrated that the male gender, higher education background, urban dwellers, government employees were significantly associated with outward bound medical tourism. This is consistent with other studies which show that in addition to other factors, age and higher education are associated with choice of health facility for treatment of various conditions [38–40]. It is also not surprising that urban dwellers are associated with medical tourism, as 27% of the urban population are insured compared to the 12% of the rural population, implying the former have greater access to finances.

Upon subjecting the data to regression analysis we found that the sociodemographic factors in used our study were not independent factors for seeking treatment in India, with respect to cancer. Our study revealed that monthly income greater than US$ 250 (OR 39) was an independent factor. This implies that initial significant sociodemographic factors were possibly indicators of increased access to required resources rather being independent influencers in the case of cancer therapy.

Other factors that influenced choice of treatment center in Kenya or India, included; perception that the country can provide required cancer treatment, availability of adequate treatment facilities, waiting time, advice from physician and friends or relatives, opinion of other patients, perceived quality of care[41], availability of specialized health care facilities, combining treatment with sight-seeing and business and cost-effectiveness of treatment. This is finding is comparable with other studies from other countries [42,43].

When we scrutinized our data further for strength of association using logistic regression, we found that like other studies the independent predicators in relation to cancer included; longer duration from diagnosis (1.16) times with every additional month), advice from physicians (OR 66 times), opinion of friends and family (OR 42) and anticipated better quality of care at chosen facility (OR 22.5) [5,19,33,42,44–48]. Similar to our results, Crush and Chikanda [37] in their editorial article demonstrated the two key reasons why patients from African Countries sought treatment in South Africa were: recommendations by their physicians and non-availability of required medical treatment. This implies a significant proportion of cancer patients who seek treatment abroad are discontent with their own home health system like in other countries [49].

All in all, we found that cost effectiveness was a significant factor for 76% of the respondents treated in India and 72% of those treated in Kenya. This is one factor the is constantly cited in most literature as a key factor for medical tourism[33,49–52]. In our study, we found that cost was an important factor for both groups, those treated in Kenya and those in India. So there was no significant difference between the two groups. However, we also found that private hospitals referred a higher proportion of respondents to India, 59 (73.8 %), than public facilities, 8 (5.6%) patients, indicating the high cost of out of care is an important ‘push’ factor [53]. Therefore, most of the respondents in our study, probably chose their facilities because they were more likely to get cheaper cancer care than the Kenyan private sector hospitals.

Several other studies have demonstrated that long waiting periods, online information, marketing, opportunity to combine sightseeing with treatment and distance were important influencing factors for medical tourism [1,7,54]. These factors did not out stand out as independent factors in our study. It is possibly due to several factors such as, marketing/promotion of healthcare is controlled and regulated [55]. Second, in the recent past, there have been concerted efforts at improving the healthcare infrastructure for cancer care with the installation of linear accelerators for radiotherapy [4], which in turn has reduced waiting time and long queues. Third, most respondents required chemotherapy for reproductive health cancers, a mode of therapy which is available in Kenya’s tertiary health facilities, like KNH. Fourth, our study focused on cancer therapy only at the exclusion of other conditions such as cardiovascular and renal disease. Investments and improvements into the health sector for management of these conditions is still lagging behind. Medical tourism is contingent on the capacity in home country to provide specialized health care with key resources being, highly skilled medics and advanced medical equipment [2].

In light of the findings of this study, and in a bid to transform Kenya and reverse outward bound medical tourism for cancer patients, the country will need address several elements to strengthening provision of specialized cancer care. Key among the elements, including: training and availing highly skilled health workers; continually upgrading the services to offer cutting edge procedures such as bone-marrow transplants; and offer cost effective packages for cost containment. Finally, it will be important to continually sensitize physicians, in private and public health facilities, and the general public on the availability of comprehensive, specialized and cost effective cancer care within the country.

### Strengths and Limitations of the Study

The study derived its data from patients themselves and not unverified media reports or private consultancies [1], making the it authentic. Further, we were able to compare those who travelled and those who were treated in Kenya from centrally and focused sites which had large enough numbers to allow for randomization.

There were a number of limitations to this study. Foremost, it was conducted within a short period of time, due to resource constraints and used recalled data which is known to introduce limitations in terms of accuracy due to recall bias in specific areas such as cost. Determining and analyzing the cost of care was not possible because of lack of verifiable documents and the use of recall bias, and as such further studies on the same are recommended.

### Recommendations for Future Research

There will be need for additional research to study other probable influencing factors such as customer care, hospital accreditation, language, climate, expected long-term outcomes of care, attitude of host country citizenry, religious accessibility and food, demonstrated in other studies [5,19]. In-depth qualitative studies are also likely to provide additional information such as patient related factors and perception of quality of care. Additional studies on physicians’ motivation factors for referrals abroad, cost-effectiveness, cost analysis, long term outcomes, impact on health systems will also be important to conduct and contribute to the body of knowledge.

## Conclusion

Medical tourism in Kenya is a growing and important phenomenon making it imperative to seek evidence-based recommendations on several issues, including patient-related motivation factors. The study identified key independent predicators of outward-bound travel that will contribute to providing evidence-based recommendations for reversing the medical tourism ‘tide’, and establishing competitive access to specialized quality care regionally while protecting health system. The key factors that influence outward bound medical tourism from Kenya are increasing duration in months from diagnosis, higher monthly income, advice from physicians, opinion of friends and family, perception of quality of care to be received, lack of adequate cancer treatment services at their local facility, and anticipation to receive better quality of care at chosen facility.

## Data Availability

All data produced in the present study are available upon reasonable request to the authors

## Abbreviations

KNH: Kenyatta National Hospital
MOH: Ministry of Health
MT: Medical Tourism
NHIF: National Hospital Insurance Fund
OR: Odds Ratio
TCC: Texas Cancer Center
QOC: Quality of Care
US$: United States Dollars.

## Acknowledgements

The authors are grateful to the late oncologist Dr Eliud Njuguna, Carol Bisieri and all the research assistants. The cooperation and support of the Ministry of Health and the management and patients of Kenyatta National Hospital and Texas Cancer Center.

## Declarations

### Consent for publication

Not applicable

### Availability of data and materials

All data generated or analysed during this study are included in this published article [and its supplementary information files].

### Authors’ contributions

MW, being the principal investigator, designed and carried out the study. MW wrote the manuscript, with input from FW, EW and PW. FN, CN and JK are content experts provided invaluable input on the research carried out. FW and EW supervised the data collection and performed the data entry. All authors read and approved the final manuscript.

## References

1. Hanefeld J, Horsfall D, Lunt N, Smith R. Medical Tourism: A Cost or Benefit to the NHS? PLoS One. 2013;8(10):1–8. doi:10.1371/journal.pone.0070406

2. Orekoya IO, Oduyoye OO. Implications of Outbound Medical Tourism on Public Health Care Development in Nigeria. Eur Sci Journal, ESJ [Internet]. 2018;14(30):353. Available from: http://dx.doi.org/10.19044/esj.2018.v14n30p353

3. Epundu U, Adinma E, Ogbonna B, Epundu O. Medical Tourism, Public Health and Economic Development in Nigeria: Issues and Prospects. Asian J Med Heal [Internet]. 2017;7(2):1–10. Available from: doi:10.9734/AJMAH/2017/36658.

4. Ministry of Health National Cancer Control Strategy 2017-2022. National Cancer Control Strategy 2017-2022. 2017. 1–76 p. Available from: http://www.health.go.ke/wp-content/uploads/2017/10/NATIONAL-CANCER-CONTROL-STRATEGY-2017-2022-KENYA-.pdf

5. Alsharif MJ, Labonté R, Zuxun Lu Z. Patients beyond borders: A study of medical tourists in four countries. Glob Soc Policy [Internet]. 2010 Dec;10(3):315–35. Available from: http://journals.sagepub.com/doi/10.1177/1468018110380003

6. Folinas S, Obeta MU, Etim GU, Etukudoh SN. COVID-19 Pandemic: The Medical Tourism and its Attendant Outcome for Nigeria. J Bus Manag 2021;23(7):26–35. Available from: https://www.iosrjournals.org/iosr-jbm/papers/Vol23-issue7/Series-6/D2307062635.pdf

7. Pocock NS, Phua KH. Medical tourism and policy implications for health systems: a conceptual framework from a comparative study of Thailand, Singapore and Malaysia. Global Health [Internet]. 2011];7:12. Available from: http://images.biomedsearch.com/21539751/1744-8603-7-12.pdf?AWSAccessKeyId=AKIAIBOKHYOLP4MBMRGQ&Expires=1493164800&Signature=5BmWL8SATpvhEvceC5DWxrRVRvk%3D

8. Yong D, Toleman MA, Giske CG, Cho HS, Sundman K, Lee K, et al. Characterization of a New Metallo--Lactamase Gene, blaNDM-1, and a Novel Erythromycin Esterase Gene Carried on a Unique Genetic Structure in Klebsiella pneumoniae Sequence Type 14 from India. Antimicrob Agents Chemother [Internet]. 2009 Dec 1;53(12):5046–54. Available from: http://www.ncbi.nlm.nih.gov/pubmed/19770275

9. Nurjadi D, Friedrich-Jänicke B, Schäfer J, Genderen PJJ, Goorhuis A, Perignon A et al. Skin and soft tissue infections in intercontinental travellers and the import of multi-resistant Staphylococcus aureus to Europe. Clin Microbiol Infect]. 2015;21(6):567. Available from: https://pubmed.ncbi.nlm.nih.gov/25753191/

10. Wangai FK, Masika MM, Maritim MC SR. Methicillin-resistant Staphylococcus aureus (MRSA) in East Africa: Red alert or red herring? BMC Infect Dis. 2019;19(1):596. Available from: https://bmcinfectdis.biomedcentral.com/articles/10.1186/s12879-019-4245-3

11. Wangai FK, Masika MM, Lule GN, Karari EM, Maritim MC, Jaoko WG et al. Bridging antimicrobial resistance knowledge gaps: The East African perspective on a global problem. PLoS One 2019;14(2). Available from: https://journals.plos.org/plosone/article?id=10.1371/journal.pone.0212131

12. Hle C, Registrar Poon ML, Associate Consultant M, Registrar Teo JWPF, Officer S, Chan EH, et al. The perils of medical tourism: NDM-1-positive Escherichia coli causing febrile neutropenia in a medical tourist. Singapore Med J [Internet]. 2011 [cited 2017 Apr 18];52(524):299–299. Available from: http://smj.sma.org.sg/5204/5204cr1.pdf

13. Centers for Disease Control and Prevention (CDC). (2010) MMWR Morb Mortal Wkly Rep 2010; Detection of Enterobacteriaceae Isolates Carrying Metallo-Beta-Lactamase ---United States, 2010. Detection of Enterobacteriaceae isolates carrying metallobeta-lactamase -United States. 2010 [cited 2017 Apr 24]. Available from: https://www.cdc.gov/mmwr/preview/mmwrhtml/mm5924a5.htm

14. Struelens marcstruelens MJ, Monnet DL, Magiorakos AP, Santos FO, Giesecke J. New Delhi metallo-beta-lactamase 1–producing Enterobacteriaceae: emergence and response in Europe, the European NDM-1 Survey Participants. Euro Surveill. 2010 [cited 2017 Apr 24];15(46). Available from: www.eurosurveillance.org:pii=19716.

15. Harling R, Turbitt D, Millar M, Ushiro-Lumb I, Lacey S, Xavier G, et al. Passage from India: an outbreak of hepatitis B linked to a patient who acquired infection from health care overseas. Public Health [Internet]. 2007 Oct];121(10):734–41. Available from: http://linkinghub.elsevier.com/retrieve/pii/S0033350607000935

16. Earls MR, Coleman DC, Brennan GI, Fleming T, Monecke S, Slickers P et al. Intra-Hospital, Inter-Hospital and Intercontinental Spread of ST78 MRSA From Two Neonatal Intensive Care Unit Outbreaks Established Using Whole-Genome Sequencing. Front Microbiol [Internet]. 2018;4(9):1485. Available from: https://doi.org/10.3389/fmicb.2018.01485

17. Nurjadi D, Fleck R, Lindner A, Schäfer J, Gertler M, Müller A et al. Import of community-associated, methicillin-resistant Staphylococcus aureus to Europe through skin and soft tissue infection in intercontinental travellers, 2011-2016. Clin Microbiol Infect [Internet]. 2019;25(6):739–46. Available from: https://pubmed.ncbi.nlm.nih.gov/30315958/

18. Mogaka JJO, Mashamba-Thompson TP, Tsoka-Gwegweni JM, Mupara LM. Effects of Medical Tourism on Health Systems in Africa. African J Hosp [Internet]. 2017 [cited 2017 Apr 17];6(1). Available from: www.ajhtl.com

19. Crooks V, Kingsbury P, Snyder J, Johnston R. What is known about the patient’s experience of medical tourism? A scoping review. BMC Health Serv Res. 2010;

20. https://www.globenewswire.com/en/news-release/2018/06/25/1528923/0/en/Global-Medical-Tourism-Market-Expected-to-Reach-USD-28-0-Billion-by-2024-Zion-Market-Research.html [Internet]. Available from: https://www.globenewswire.com/en/news-release/2018/06/25/1528923/0/en/Global-Medical-Tourism-Market-Expected-to-Reach-USD-28-0-Billion-by-2024-Zion-Market-Research.html

21. Mogaka JJ, Tsoka-Gwegweni JM, Mupara LM, Mashamba-Thompson T. Role, structure and effects of medical tourism in Africa: A systematic scoping review protocol. Vol. 7, BMJ Open. 2017. doi:10.1371/journal.pone.0070406

22. Republic of Kenya October 2007. Kenya Vision 2030: A globally competitive and prosperous Kenya Available from: http://www.nayakenya.org/pdf/12.pdf

23. Ministry of Health Kenya Health Policy 2014-2030 [Internet]. Ministry of Health; 2014 [cited 2017 Apr 24]. Available from: https://www.health.go.ke

24. Njuguna D, Wanjala P. A Case for Increasing Public Investments in Health. A Case Increasing Public Investments Heal [Internet]. 2019; Available from: https://www.health.go.ke/wp-content/uploads/2019/01/Healthcare-financing-Policy-Brief.pdf

25. Kenya National Bureau of Statistics. Economic Survey Report 2020 [Internet]. Economic Survey 2021. Statistics, Kenya National Bureau of; 2021. 127–128 p. Available from: https://www.knbs.or.ke/?wpdmpro=economic-survey-2020

26. Republic of Kenya Ministry of health. 2019 Kenya Health Accounts 2015/2016. Government of Kenya; 2019. Available from: www.health.go.ke

27. Ministry of Health. 2018 Kenya Household Health Expenditure and Utilization Survey. Nairobi: Government of Kenya; 2018. 133 p. Available from: http://www.health.go.ke

28. The Republic of Kenya (2010) The Constitution of Kenya Revised Ed.2010 National Council for Law Reporting, Government Printers

29. Fleiss JL, Levin B, Paik MC. Statistical methods for rates and proportions. John Wiley & Sons; 2013.

30. Sultana S, Haque A, Momen A, Yasmin F. Factors affecting the attractiveness of medical tourism destination: an empirical study on India-review article. Iran J Public Health [Internet]. 2014 Jul [cited 2019 Aug 25];43(7):867–76. Available from: http://www.ncbi.nlm.nih.gov/pubmed/25909055

31. Ruggeri K, Ruggeri K, Záliš L, Meurice CR, Hilton I, Ly T, et al. Evidence on global medical travel. Bull World Heal Organ [Internet]. 2015;(August 2014):6–17. Available from: http://www.ncbi.nlm.nih.gov/pubmed/26549906

32. Badulescu D, Badulescu A. Medical tourism: between entrepreneurship opportunities and bioethics boundaries: narrative review article. Iran J Public Health [Internet]. 2014 Apr [cited 2017 Mar 29];43(4):406–15. Available from: http://www.ncbi.nlm.nih.gov/pubmed/26005650

33. Hanefeld J, Lunt N, Smith R, Horsfall D. Why do medical tourists travel to where they do? The role of networks in determining medical travel. Soc Sci Med [Internet]. 2015 [cited 2017 Apr 3];124:356–63. Available from: http://www.sciencedirect.com/science/article/pii/S0277953614003104

34. Goldberg AM. Medical tourism? A case study of African patients in India. UC Berkeley Theses, [Internet]. 2013;15(4):250–60. Available from: https://escholarship.org/uc/item/89t9j2b8

35. Globocan. Kenya Source. 2020;799:2020–1. Available from: https://gco.iarc.fr/today/data/factsheets/populations/404-kenya-fact-sheets.pdf

36. Sung H, Ferlay J, Siegel RL, Laversanne M, Soerjomataram I, Jemal A, et al. Global Cancer Statistics 2020: GLOBOCAN Estimates of Incidence and Mortality Worldwide for 36 Cancers in 185 Countries. CA Cancer J Clin [Internet]. 2021;71(3):209–49. Available from: cacancerjournal.com

37. Crush J, Chikanda A. South to South medical tourism and the quest for health in Southern Africa. Soc Sci Med [Internet]. 2014; Available from: http://jptedsi.ir/wp-content/uploads/2015/10/South-South-medical-tourism-and-the-quest-for-health-in-Southern-Africa.pdf

38. Victoor A, Delnoij DMJ, Friele RD, Rademakers Jjdjm. Determinants of patient choice of healthcare providers: a scoping review. BMC Health Serv Res. 2012 Aug 22;12:272. Available from: http://www.ncbi.nlm.nih.gov/pubmed/22913549

39. Damman OC, Spreeuwenberg P, Rademakers J, Hendriks M. Creating compact comparative health care information: what are the key quality attributes to present for cataract and total hip or knee replacement surgery? Med Decis Mak. 2012;32(2):287–300.

40. Robertson R, Burge1 P. The impact of patient choice of provider on equity: analysis of a patient survey. J Health Serv Res Policy. 2011;16(1_suppl):22–8.

41. Berry LL, Parasuraman A, Zeithaml VA. SERVQUAL: A multiple-item scale for measuring consumer perceptions of service quality. J Retail. 1988;64(1):12–40. Available from: https://www.researchgate.net/profile/Valarie-Zeithaml-2/publication/225083802_SERVQUAL_A_multiple-_Item_Scale_for_measuring_consumer_perceptions_of_service_quality/links/5429a4540cf27e39fa8e6531/SERVQUAL-A-multiple-Item-Scale-for-measuring-consumer-percep

42. Khan MJ, Chelliah S, Haron MS, Ahmed S. Role of Travel Motivations, Perceived Risks and Travel Constraints on Destination Image and Visit Intention in Medical Tourism: Theoretical model. Sultan Qaboos Univ Med J. 2017 Feb;17(1):e11–7. Available from: http://www.ncbi.nlm.nih.gov/pubmed/28417022

43. Anish MN DP& SR. Antecedents to behavioural intentions in medical tourism. Int J Manag Decis making [Internet]. 2016;15(No.3/4). Available from: https://www.inderscienceonline.com/doi/pdf/10.1504/IJMDM.2016.080706

44. Runnels V, Carrera PM. Why do patients engage in medical tourism? Maturitas [Internet]. 2012 Dec;73(4):300–4. Available from: http://www.ncbi.nlm.nih.gov/pubmed/23007007

45. Connell J. Contemporary medical tourism: Conceptualisation, culture and commodification. Vol. 34, Tourism Management. 2013. p. 1–13.

46. Khan MJ, Chelliah S, Haron MS. International Patients’ Travel Decision Making Process-A Conceptual Framework. Iran J Public Health [Internet]. 2016 Feb [cited 2019 Aug 16];45(2):134–45. Available from: http://www.ncbi.nlm.nih.gov/pubmed/27114978

47. Yeoh E, Othman K, Ahmad H. Understanding medical tourists: Word-of-mouth and viral marketing as potent marketing tools. Tour Manag. 2013 Feb;34:196–201. Available from: https://linkinghub.elsevier.com/retrieve/pii/S026151771200088X

48. Helble M. The Movement of Patients across borders:challenges and opportunities for public health. Bull World Health Organ. 2011;89(1):68–72.

49. Hanefeld J, Smith R, Horsfall D, Lunt N. What do we know about medical tourism? A review of the literature with discussion of its implications for the UK national health service as an example of a public health care system. J Travel Med. 2014;21(6):410–7.

50. Ruggeri K, Záliš L, Meurice CR, Hilton I, Ly TL, Zupan Z, et al. Evidence on global medical travel. Bull World Health Organ [Internet]. 2015;93(11):785–9. Available from: http://www.ncbi.nlm.nih.gov/pubmed/26549906

51. Paul DP, Barker T, Watts AL, Messinger A, Coustasse A. Insurance Companies Adapting to Trends by Adopting Medical Tourism. Health Care Manag. 2017;36(4):326–33. Available from: http://www.ncbi.nlm.nih.gov/pubmed/28953068

52. Fisher C, Sood K. What Is Driving the Growth in Medical Tourism? Health Mark Q [Internet]. 2014 Jul 3;31(3):246–62. Available from: http://www.ncbi.nlm.nih.gov/pubmed/25120045

53. Noree T, Hanefled J, Smith R. Medical Tourism in Thailand: a cross sectional study [Internet]. Vol. 94, Bulletin of the World Health Organization. 2016. p. 30–6. Available from: http://europepmc.org/abstract/MED/26769994

54. Ahwireng-Obeng F van LC. Africa’s middle class women bring entrepreneurial opportunities in breast care medical tourism to South Africa. Int J Heal Plann Manag. 2011;(26):39–55.

55. Republic of Kenya. The Medical Practitioners and Dentists (Practitioners and Health Facilities) (Advertising) Rules, 2016. Kenya Subsid Legis Leg Not 130. 2016;

